# Tentative analysis of biomarkers associated with filariasis in moungo division, littoral, cameroon

**DOI:** 10.1101/2024.08.29.24312770

**Authors:** Jean Baptiste Hzounda Fokou, Syntiche Teudem Biyong, Francine Kouemo, Fru Awah Akumwah, Ambassa Reine, Véronique Simone Fannang, Juliette Koube, Jules Clement Assob

## Abstract

Filariasis is a significant cause of morbidity and a public health concern in most tropical countries. These diseases are usually contracted in childhood and most often diagnosed in adulthood. This study aimed to identify biochemical markers associated with filariasis and the endobacteria present in microfilariae.

This was a cross-sectional analytical study within a hospital setting. Persons aged from six years and above were included in the study. They were then clustered into asymptomatic and symptomatic. Data was collected from February to June 2023. Blood, urine, and skin samples were collected and analysed by microscopy to identify microfilariae. The plasmatic and urinary markers were evaluated by spectrophotometry and protein fraction was monitored by electrophoresis. The data was analysed using SPSS 25 with the significant threshold set at 0.005.

A total of 55 persons were included in the study, representing 74,32% of the total population questioned. The sex ratio (F/M) was 2.66. The infection rate was 18.2% *Onchocerca volvulus*, 54.54% *Mansonella perstans*, 22.72% *Loa loa* and 4.54% *Wuchereria bancrofti*. Protein levels were 35% higher in the asymptomatic with no parasite, 80% in the asymptomatic with detected parasite and 91.67% in the symptomatic. The decrease in albumin fraction and increase in gamma fractions seem to be correlated with the presence of parasites in the blood.

These results suggest that protidemia and serum protein are significant indicators in the diagnosis of filariasis.

## Introduction

Filariasis is defined by The World Health Organisation (WHO) as a parasitic disease caused by the presence of filarial nematodes and their embryos in the body, microfilariae, which for a long time was classified as a "neglected" tropical disease [1,2]. Filariasis is a group of diseases that vary according to the parasite species, the clinical manifestations and the nature of the immune response induced [3]. Their socio-economic impact makes them a major cause of morbidity in most tropical countries [4–10].

It is estimated that over 200 million people worldwide are affected by filariasis out of approximately one billion people at risk [1]. Current global estimates suggest that by 2019, 863 million people in 47 countries will be at risk of lymphatic filariasis, with 30% of these people living in the African region [9;12;13]. The number of persons infected with onchocerciasis is estimated at around 18 million, with over 99% living in intertropical Africa [13]. In Central Africa, 15 to 20 million people are infected with schistosomiasis and more than 50 million people are infected with dracunculiasis (Medina filaria) [14]. Several outbreaks have been described in Cameroon, notably in the town of Nkongsamba since 1937 [5]. According to the distribution of endemicity of neglected tropical diseases in Cameroon in 2017, 152 districts are endemic for lymphatic filariasis, 113 districts are endemic for onchocerciasis, and the prevalence of co-endemicity for schistosomiasis ranges from 0.2% to 6.2% [15].

Over the past few years, several therapeutic strategies have been developed, yet mass treatment is not recommended in regions where there is co-endemicity with loiasis, a type of filariasis that is just as challenging as onchocerciasis and volvulus [1,16,17]. Low-specific antigenic tests and Loa-scop have been developed for the diagnosis of filariasis, yet remain inaccessible in most endemic regions. The selection of populations infected with onchocerciasis or loiasis, respectively, means that they can be treated with ivermectin (Mectizan®), without the risk of developing a serious adverse reaction requiring palliative care. However, those who are not eligible for this mass treatment are given an individual long-lasting treatment [19;20]. Moreover, the risk of resistant microfilariae is a major concern, and the ineffectiveness of available antiparasitic drugs on adult worms represents a significant impairment to breaking the chain of transmission [20].

Filariasis is typically contracted during early childhood, with the majority of cases presenting with no symptoms. However, acute and chronic symptoms may subsequently emerge in adulthood [9]. Therefore, it is of the utmost importance to identify specific markers for filariasis, which could pave the way for the development of an early diagnosis method, the creation of a novel treatment, or even a vaccine. To achieve this, it is first necessary to conduct a preliminary mapping of all the metabolites that are directly or indirectly involved in the pathogenicity of the different forms of filariasis. The objective of this study is to identify biochemical markers associated with filariasis and the metabolome of the endobacteria present in the microfilariae identified.

## Material and Methods

### Type of study

A cross-sectional and analytical study was conducted.

### Study site

Specimens were collected in public and private hospitals in the Moungo Division, with particular focus on the Nkongsamba Regional Hospital (NRH), the Ékol-Mbeng Integrated Health Centre (CSI-Ékol-mbeng), the Manengolè Catholic Medical Health Centre (CSMCM) and the Bon Samaritain private health centre in Njombé.

The samples were analysed at the Laboratory of Pharmacology, Faculty of Medicine and Pharmaceutical Sciences at the University of Douala, at Douala General Hospital and in the bacteriology laboratory at the Centre Pasteur du Cameroun in Yaoundé.

### The study population

The study population consisted of patients of any sex aged at least six years.

### Inclusion criteria

Any person of any sex, aged at least 6 years, who consented and who visited the hospitals concerned during the study period was included in the study.

### Non-inclusion criteria

The following individuals were not selected for this study: hospital patients and children under the age of six.

### Exclusion criteria

The study excluded three categories of participants: pregnant women; volunteers who had withdrawn their informed consent; and any individual with HIV, AIDS, or cancer, or another serious disease requiring specific monitoring, due to the potential for bias in our results caused by other diseases present.

### Sampling

The sampling was non-exhaustive and consecutive, and the sample size was determined by the following Cochran formula.

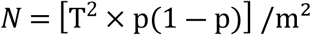

with

*N: The size of the sample*

*T: The standard deviation for the normal distribution, with a 95% confidence interval of 1.96*

*p: The prevalence of filariasis*

*m: The margin of error at 10%*

*Standard value for the margin of error: 0.1*

*The prevalence of filariasis in Cameroon is estimated to be 17%* [21].

*Consequently, N can be calculated as follows:*

*N = [1.96² × 0.17 (1-0.17)] / 0.1² = 54.20*

*Consequently, the minimum sample size was set at 54 participants*.

## Technical procedures

### Patient recruitment

Some patients were contacted by telephone, while others were approached and recruited in the consultation and sampling units of the laboratory departments of the hospitals involved in the study. Prior to their participation, all patients were given a detailed verbal presentation of the aim of the study. For those who agreed to take part, an informed consent form was provided for their signature, along with a data collection form and they were assisted when needed in filing the form.

### Data Collection

Data were collected using an individual survey form (adults and minors). Data were collected from interviews with patients and/or parents or guardians who had read, understood and consented to participate in the study after the purpose of the study had been explained.

*The following variables were collected:*

- Socio-demographic data (age, sex, occupation, place and duration of residence in the region)
- Epidemiological and clinical data
- Biological data

### Pre-analytical steps

#### Blood sampling

Venipuncture was done at the elbow or back of the hand during the daytime (12:00-16:00) in a plain tube for the determination of plasma markers (PCT, Total Proteins, Electrophoresis). An EDTA tube was used for the detection of *Loa loa* and *Mansonella perstans* microfilariae (thick film, leuco-concentration), and a simple prick at the fingertip was employed to collect the first drop of blood (fresh state) when possible. The nightly sampling period (22:00-01:00) entailed a simple venipuncture procedure at the elbow or back of the hand, conducted in an EDTA tube, with the objective of testing for *Wuchereria bancrofti*.

### The sampling procedure employed was as follows

Once the required equipment has been assembled and the patient’s previously identified tubes and slides have been located, a tourniquet is applied approximately 7 to 10 cm above the venipuncture site. The patient is then asked to point the bandage so that the vein is turgid, after which the vein is palpated and the puncture site is cleaned using a cotton wool pad soaked in povidone iodine 10%, which is then disposed of in the bin. Subsequently, the needle and pump body should be assembled and the needle inserted into the vein, with the bevel facing upwards. The collection tube is then inserted into the vacutainer and filled to the mark. Subsequently, the tube was removed from the holder and a dry swab was placed over the puncture site. Subsequently, the needle was also removed and a dry pad was applied to the lesion area in order to facilitate haemostasis. Once this had been achieved, the pad was removed and a dry dressing was applied. The needle was then disconnected and discarded into the provided safety box. The sample was later secured and coded, the participant’s little finger was massaged and the fingertip was disinfected. A brief puncture was made and the first drop was collected on a slide that had been previously identified and coded. Next, a dry cotton pad was placed over the puncture site to prevent further bleeding. A slide cover was placed over the freshly collected drop of blood and observed under an optical microscope as soon as possible.

### Skin sampling

Skin sampling was conducted at the Ekol-Mbeng Centre for the Study of Infectious Diseases (CSI), also known as the "Centre des Grands Endémiques", which specialises in filarial infections, and at the Nkongsamba Regional Hospital.

Following disinfection of the sampling area, a sterile needle was employed to pinch a flap of skin. This was then incised with a scalpel blade and placed on a slide with a drop of physiological water between the slide and the slide covers. The flap was observed twice under a light microscope, with an interval of 60 minutes between observations.

### Urine collection

Following the labelling and coding of the urine jars, the participants were provided with instructions on how to collect the urine. The urine was collected by the participants themselves in the toilets of the hospitals.

### Transport and storage

Following sampling, the samples collected were transported to the parasitology and biochemistry laboratories. Samples from dry tubes were centrifuged at 3500 rpm for five (05) minutes, and the sera obtained were aliquoted and stored at -20°C until the day of analysis. Serums in EDTA tubes were stored at room temperature for three weeks before analysis.

Finally, the urine samples were analysed without delay in the biochemistry laboratories of the various hospitals where the samples were taken.

### Analytical stage

The samples were analysed as follows:

### Microscopy

Identification, quantification and isolation of microfilariae.

### Identification and quantification of microfilariae - Giemsa staining of the thick film (GE) and blood smear

*Principle:* The Giemsa solution is a mixture of eosin and methylene blue (azure). Methylene blue colours the parasite cytoplasm in blue. The ideal pH for highlighting the granulations associated with the parasites and identifying the different species is 7.2. Several methods are used to stain EG/blood smears. For this study, we opted for the slow method, which involves preparing a 3% Giemsa dye solution for staining times of 45-60 minutes.

### Procedure

After spreading the drop of blood on a slide and drawing the smear, allow to air dry, then fix the smear by dipping it for two seconds in a jar containing methanol, then allow to air dry. Place the slides on the staining rack making sure that they do not touch each other, then carefully pour the staining solution over each slide until the spreads are covered. Allow to stain for 45 minutes, then carefully remove the remaining stain and gently rinse the slides in a jar of water buffered at pH 7.2. Then allow the slides to air dry and discard the remaining Giemsa solution. The prepared slides were read at objective 10 and with a drop of immersion oil at objective 100 [42].

Observations: Presence of a head, tail, sheath (optional), one or two internal bodies and somatic nuclei (purple).

### Isolation of microfilariae by haemolysis

Principle: This consists of destroying the red blood cells contained in a whole blood sample to better identify the microfilariae.

Reagents: 2% saponin solution (pure saponin (S 7900 Sigma® 0.2g) 0.9% NaCl solution.

In a Falcons-type centrifuge tube of at least 20 ml, place 5 ml of blood taken on anticoagulant (or 1 volume). Add 10 ml of NaCl 0.9% (or two volumes) and turn to mix well. Then add the saponin solution dropwise (250 µL). Close and invert the tube several times to ensure complete haemolysis, particularly at the end cone of the tube, as the liquid should be completely translucent. Centrifuge at 500g (approximately 1,500 rpm) for 10 minutes and discard the supernatant, without inverting the tube completely, by wiping the sides with cotton wool or sponge paper mounted on forceps so that the haemolysis liquid does not dilute the pellet when it is turned upside down. Add one drop of NaCl 0.9%. Homogenise the pellet well, then take 20 µL for quantification by the calibrated GE, and keep the rest for bacteriological and spectrometric analysis [22].

### Evaluation of plasma and urinary biochemical markers

#### Determination of Procalcitonin (PCT)

***Principle:*** Finecare PCT rapid quantitative tests are based on fluorescence immunoassay technology for the quantitative determination of PCT concentrations in whole blood, serum or plasma. Add the sample to the intended sample dilution solution and mix; then add the mixed sample to the sample hole of the corresponding test cartridge, where it will combine with the fluorescently labelled PCT antibody in the labelling buffer to form reaction complexes. As the compounds migrate onto the nitrocellulose matrix by capillary action, they are captured by the PCT antibodies that have been immobilised on the detection line of the nitrocellulose membrane. The more PCT is present in the sample, the more the complexes accumulate on the detection line. The intensity of the PCT antibody detection signal is proportional to the concentration of PCT in the specimen. The procedure strictly follows the manufacturer’s instructions.

### Evaluation of the urine strip with the URS-11 kit

The literature review informs us that in certain patients with a high parasite load, microfilariae may be found in the urine, and their presence in the urinary tract could not be the cause of certain urinary tract infections in the patient [3]. We evaluated three (03) of the eleven parameters available in the URS-11 kit. The procedure was as describe by the manufacturer.

### Determination of total protein using the Mindray BA-80 semi-automatic spectrophotometer

Principle: The Mindray BA-88A semi-automatic analyser is a spectrophotometer used for the quantitative determination of many biochemical tests. Its principle is based on spectrophotometry by measuring the absorbance of a solution at a given wavelength according to Beer Lambert’s law, which stipulates that the absorbance of a solution is proportional to the concentration of the element it contains.

### Serum protein electrophoresis on the MINICAP® instrument Serum protein electrophoresis is a technique used to separate proteins into fractions of different mobility, obtaining their relative percentages

Principle: The MINICAP system uses the principle of capillary electrophoresis in free solution, allowing the separation of charged molecules according to their electrophoretic mobility in a given pH buffer, and depending on the pH of the electrolyte, a greater or lesser electroosmotic flow. The MINICAP system comprises 2 capillaries in parallel, allowing 2 simultaneous analyses. With this system, the sample (diluted in the analysis buffer) is injected into the capillaries by suction at the anode. Separation is then achieved by applying a potential difference of several thousand volts across each capillary, and the different fractions migrate according to their electrophoretic mobilities.

The direct detection of proteins is carried out by absorption spectrophotometry spectrophotometry at 200 nm on the cathode side. The capillaries are then washed with a washing solution, followed by the analysis buffer. Interpretation of protein electrophoresis is made by relating each fraction to the level of total serum protein (g/l). With the buffer used at basic pH (9.9), the order of migration of proteins with their serum constituents is as follows gamma globulins (7.5 to 16 g/L); beta-2 globulins (containing transferrin, fibrinogen transferrin, fibrinogen and the C3 fraction of complement); beta-1 globulins; alpha-2 globulins (6 to 10 g/L) consisting in particular of alpha-2 -macroglobulin, haptoglobin; alpha-1 globulins (1.5 to 4 g/L) including alpha-1 -antitrypsin, alpha-1 -anti chymotrypsin and orosomucoid; and albumin (33 to 50 g/L).

### Statistical analysis

Microsoft Office Excel 2016 was used to enter our data; SPSS 25 for Windows (Statistical Package for Social Sciences) was used to analyse the data; Quantitative variables were represented as histograms and categorical variables as pie charts;

The Chi-square test was used to compare discontinuous quantitative variables (frequencies, proportions); and the Student t-test for continuous quantitative variables (mean and standard deviation); The difference was considered statistically significant for a value of p<0.05.

### Ethical considerations

In order to comply with the rules of biomedical ethics, we requested and obtained:

- Ethical clearance from the institutional Ethical committee of the University of Douala (3586/CEI-Udo/06/2023/M.)

Research authorization from:

- Regional Hospital of Nkongsamba (072/AR/MINSANTE/DRSPL/HRN/23)

- Njombe-Pendja health district (148/23MINSANTE/DRSPL/DSNP)

Ekol-mbeng health centre (001/L/MS/DRSL/SSDN/CSIE)

Informed consent from the participants, which consisted of having them read and understand the purpose of the study from top to bottom.

The study was conducted following good clinical laboratory practice. The anonymity of the participants and the confidentiality of the data obtained from the survey questionnaires were respected, and the results of the analyses were used exclusively for research purposes. Participation in the study was strictly voluntary.

## RESULTS

### Sociodemographic characteristics and clinical history of the population

#### Sociodemographic characteristics of the population

Our population was predominated by female (73%), they represented around 1/3 of our study population (sex ratio F/H of 2.66). The mean age of the population was 35 ± 2 years with extremes of 6 and 71 years.

### Distribution of the population according to collection sites, location, lifestyle, knowledge of the disease and sampling periods

29.09% of our population was sampled at the HRN, 20% at the Ekole Mbeng CSI, 23.63% at the CSCM and 27.27% at the private health centres of Njombé-Penja. The majority came from Nkongsamba, Manengolè, Njombé and Mboébo, with a higher distribution in Nkongsamba. Most (54%) had been born and raised in these localities, and 24% had been living there for more than 10 years. 51% of our population were working in the rural area, and 31% were still scholars. Nearly 80% of the population had knowledge of filariasis and 25% did not sleep under an impregnated mosquito net as recommended. 52.72% agreed to have their samples taken during the two sampling periods, and the rest of the participants agreed to just one sampling period (Table 1).

**Table 1:**
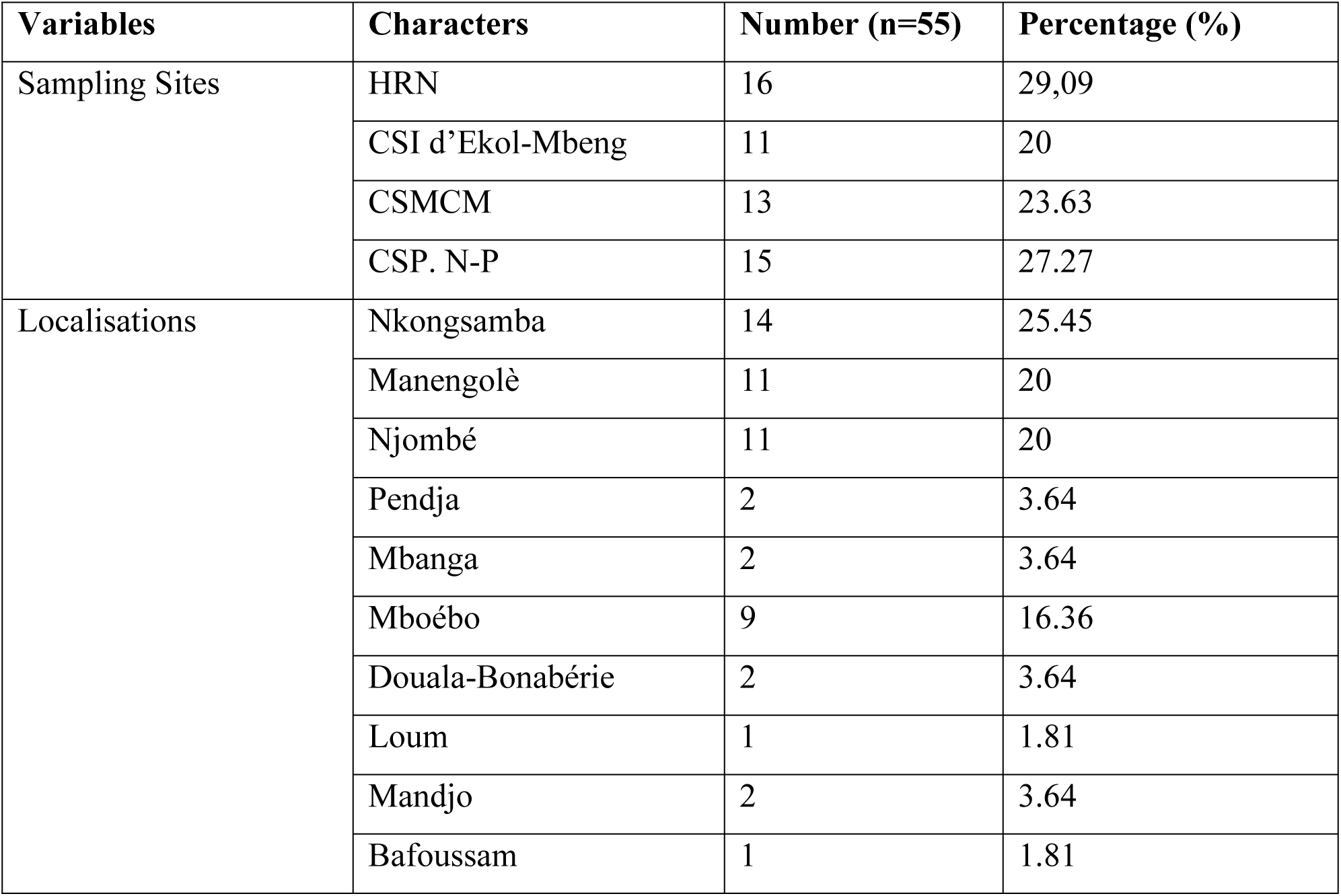
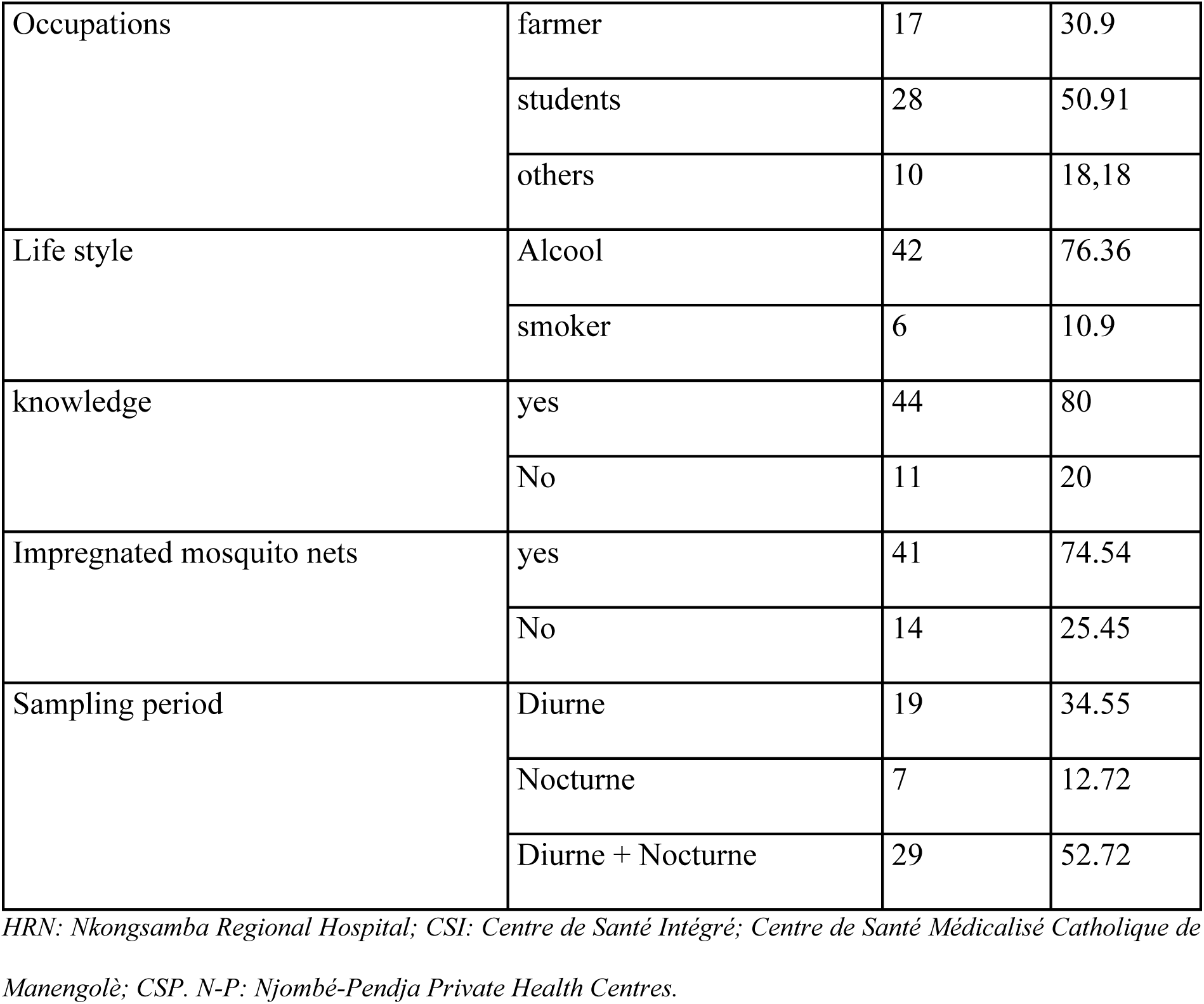
Patient distribution and lifestyle.

### Clinical history of the population

#### Distribution of the population according to the administration of mass treatment

9.09% of the population had never taken the mass treatment offered free of charge by the government, due to their fear of developing serious and disfiguring side effects. As the Mectizan® distribution campaign is carried out once a year, we noted during this study that people who had taken the treatment 1 year ago were statistically more numerous than others, as summarised in the following figure 1.

**Figure 1:**
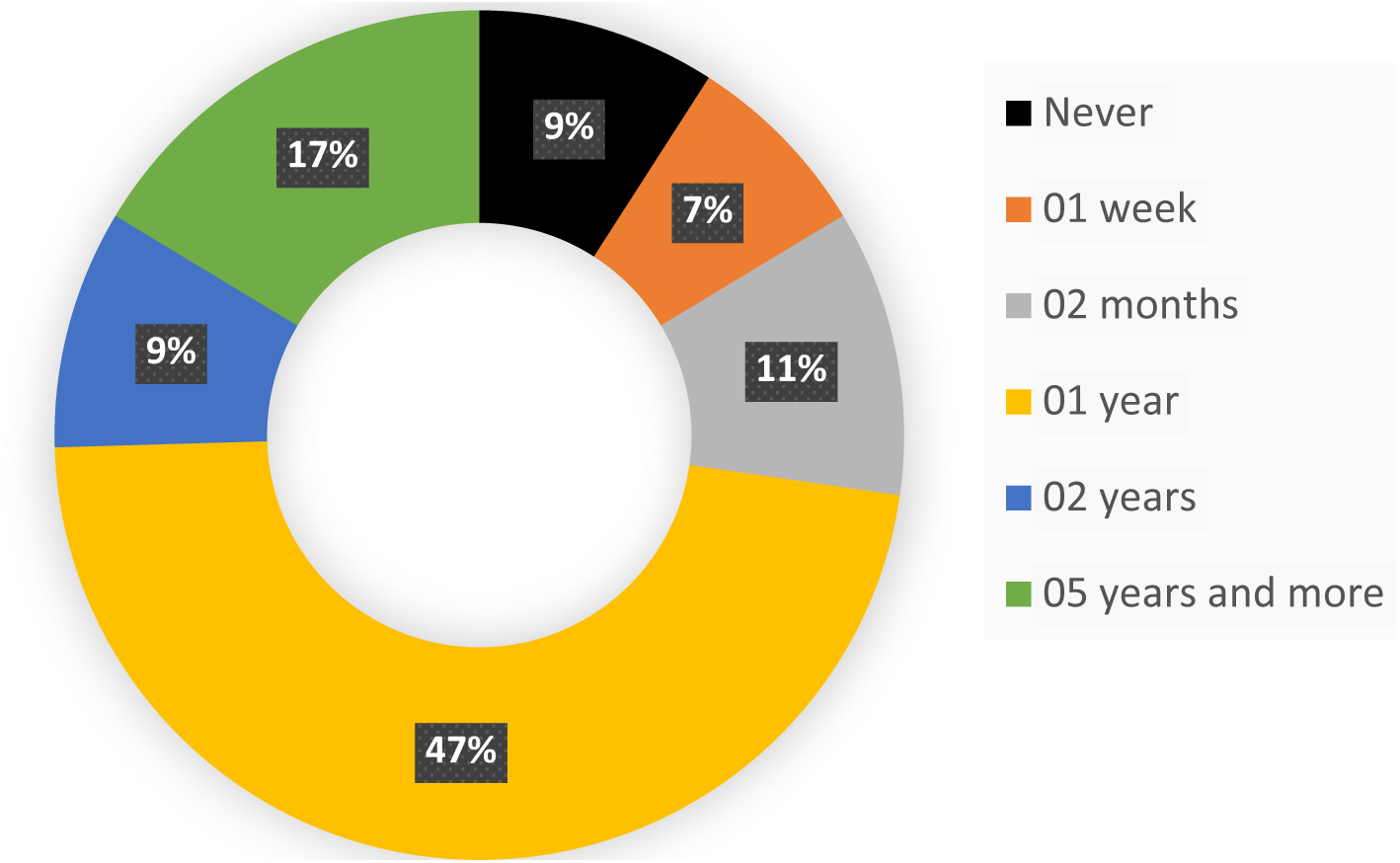
participants distribution according to Mectizan® treatment.

### Distribution of the population according to the presence or absence of symptoms

Figure 2 shows that of the 55 participants selected for our study, 49% had no symptoms at the time of sampling, while the other 51% had some symptoms of filariasis, in particular elephantiasis of the lower limbs, subcutaneous nodules, itching on the hands and buttocks, etc.

**Figure 2:**
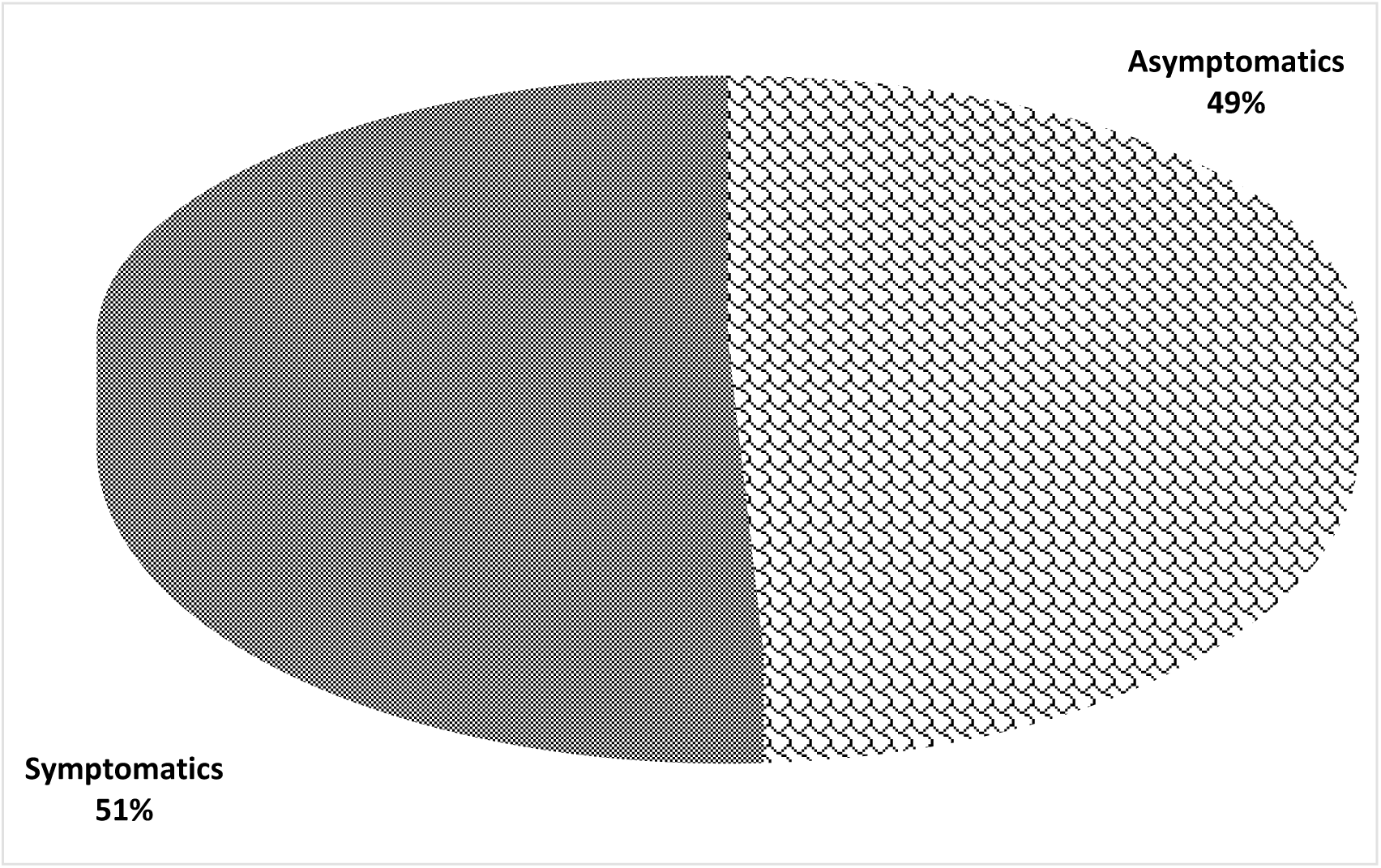
Repartition according to the symptomatology

### Identification of microfilariae

#### Breakdown of participants into groups of Symptomatic (S) and Asymptomatic (AS)

Out of 27 asymptomatic participants, 18.18% were positive cases, i.e. infected but asymptomatic patients, and out of 28 symptomatic participants, 21.81% were positive cases, i.e. infected and symptomatic patients, as summarised in Figure 3.

**Figure 3:**
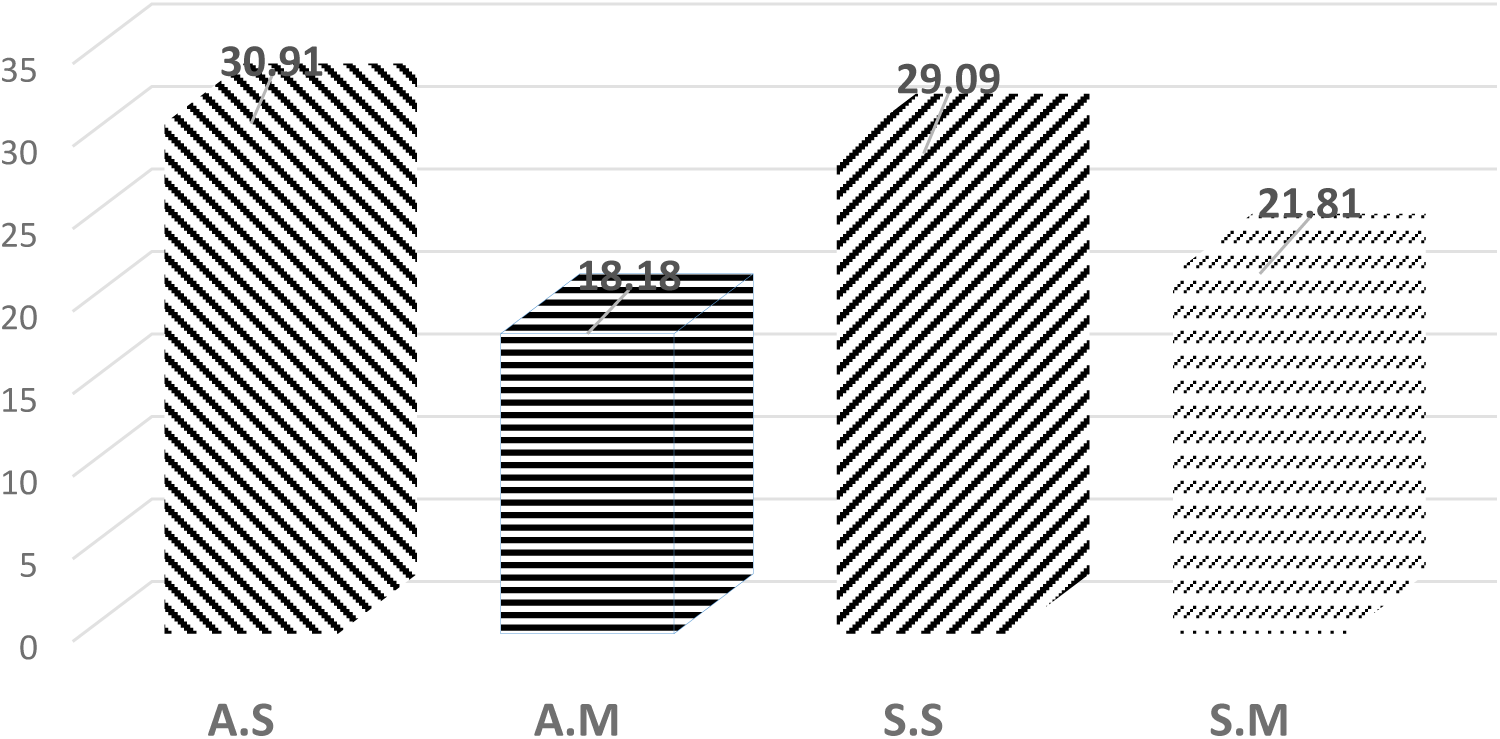
Distribution of symptomatic and asymptomatic participants

**Figure 4:**
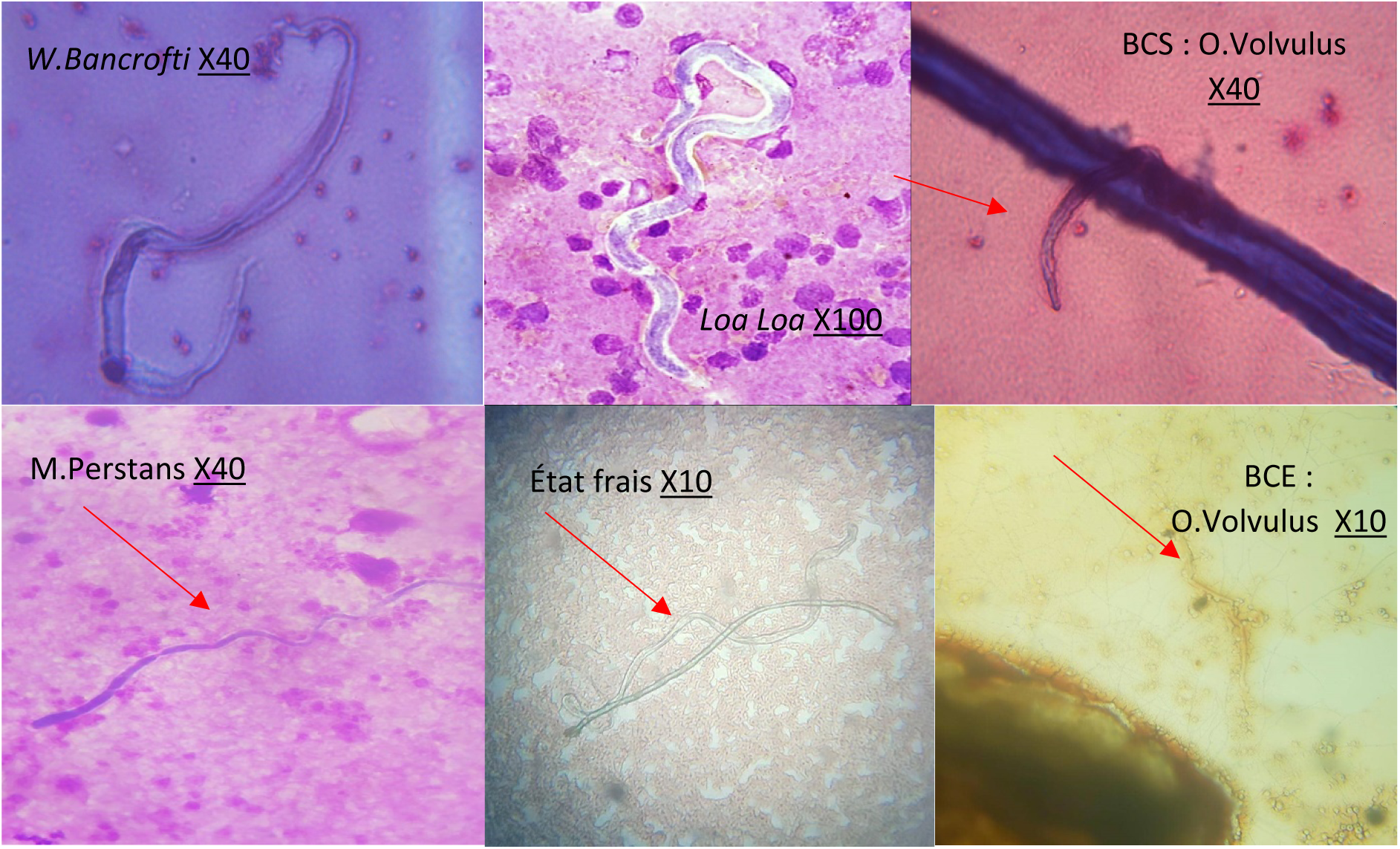
Microfilariae observed under the light microscope.

### Identification of microfilariae and distribution according to type of sample and infection

A total of 22 samples tested positive for microfilariae, present in mono infection and/or co-infection with another type of microfilaria. We recorded 01 co-infection out of 10 in asymptomatic patients and 5 co-infections out of 12 in symptomatic patients. This is summarised in Table III below.

**Table III:**
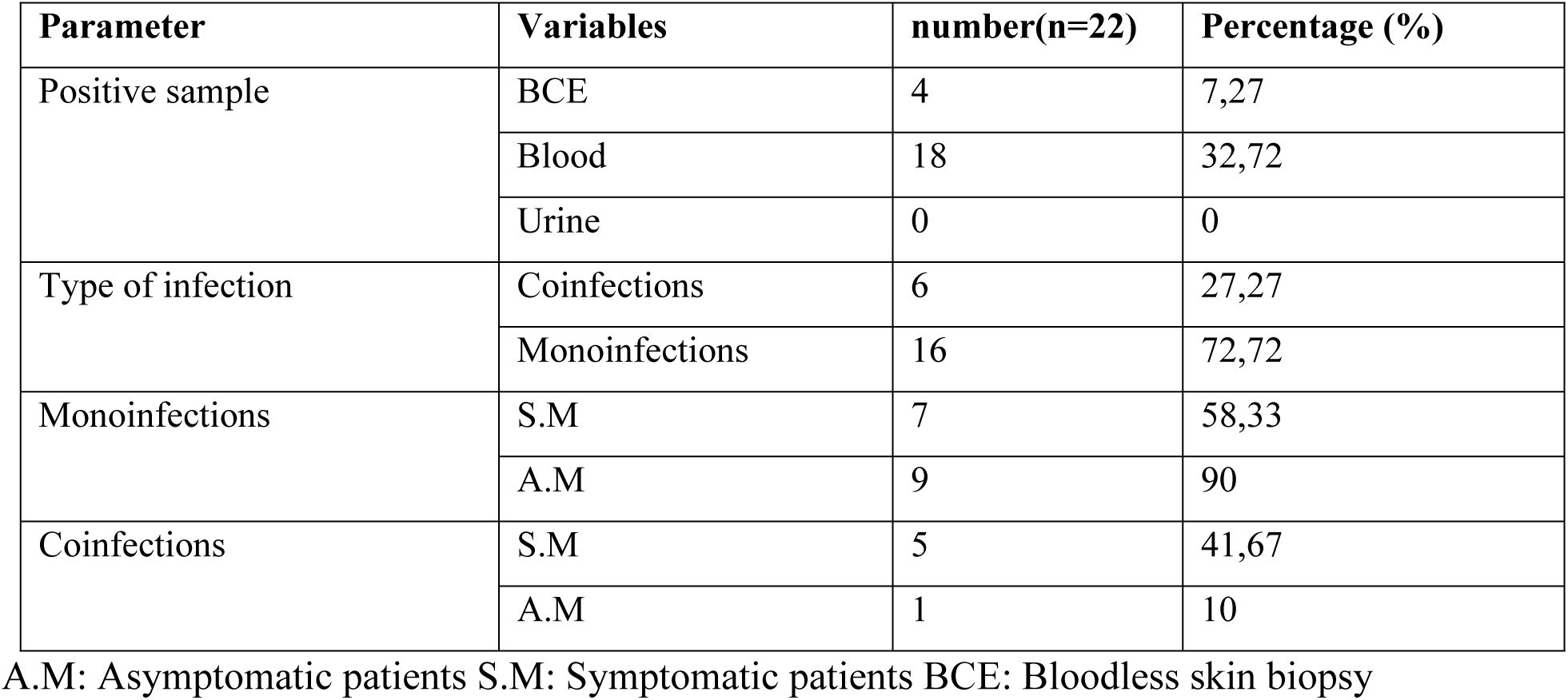
Identification and distribution of microfilariae.

| Parameter | Variables | number(n=22) | Percentage (%) |
| --- | --- | --- | --- |
| Positive sample | BCE | 4 | 7,27 |
|  | Blood | 18 | 32,72 |
|  | Urine | 0 | 0 |
| Type of infection | Coinfections | 6 | 27,27 |
|  | Monoinfections | 16 | 72,72 |
| Monoinfections | S.M | 7 | 58,33 |
|  | A.M | 9 | 90 |
| Coinfections | S.M | 5 | 41,67 |
|  | A.M | 1 | 10 |

A.M: Asymptomatic patients S.M: Symptomatic patients BCE: Bloodless skin biopsy

### Microfilariae identified and parasitic loads

*Onchocerca volvulus* was identified in 4 patients (18.2%), *Wuchereria brancrofti* in a single patient (4.54%), *Loa loa* in 5 patients (22.72%) and *Mansonella perstans* in 12 patients (54.54%). Parasitic loads were a minimum of 1 mf/μL of blood, and a maximum of 4 mf/20μL of blood, as summarised in Table IV below.

**Table IV:** Microfilariae identified and parasite loads.

| Microfilaires | Cases | parasitic loads (mf/μL) |
| --- | --- | --- |
| *Onchocerca volvulus* | 4 | [1-2] |
| *Wuchereria bancrofti* | 1 | 1 |
| *Loa loa* | 5 | [1-2] |
| *Mansonella perstans* | 12 | [2-4] |

### Type of infection

33.33% of patients were co-infected with *Onchocerca volvulus* and *Mansonella perstans*, and 66.67% were co-infected with *Loa loa* and *Mansonella Perstans*. 6.25% of patients were mono-infected with *Loa loa* and *Wuchereria bancrofti* respectively, 12.5% of patients were infected with *Onchocerca volvulus*, and finally, 75% of patients were infected with *Mansonella perstans*.

### Results of biochemical parameters of participants

#### Urine dipstick and procalcitonin results

To avoid bias in the results of biochemical parameters, we subsequently excluded all symptomatic healthy patients, as the fact of presenting symptoms without any microfilaria found could be due to another pathology.

The results of the urinary biochemistry are presented in Table V. From the table, it came out that, 15.38% had proteinuria while 2.56% of patients had leukocytes in their urine. There were no positive nitrites, and no procalcitonin was detected in the urine of the participants

**Table V:**
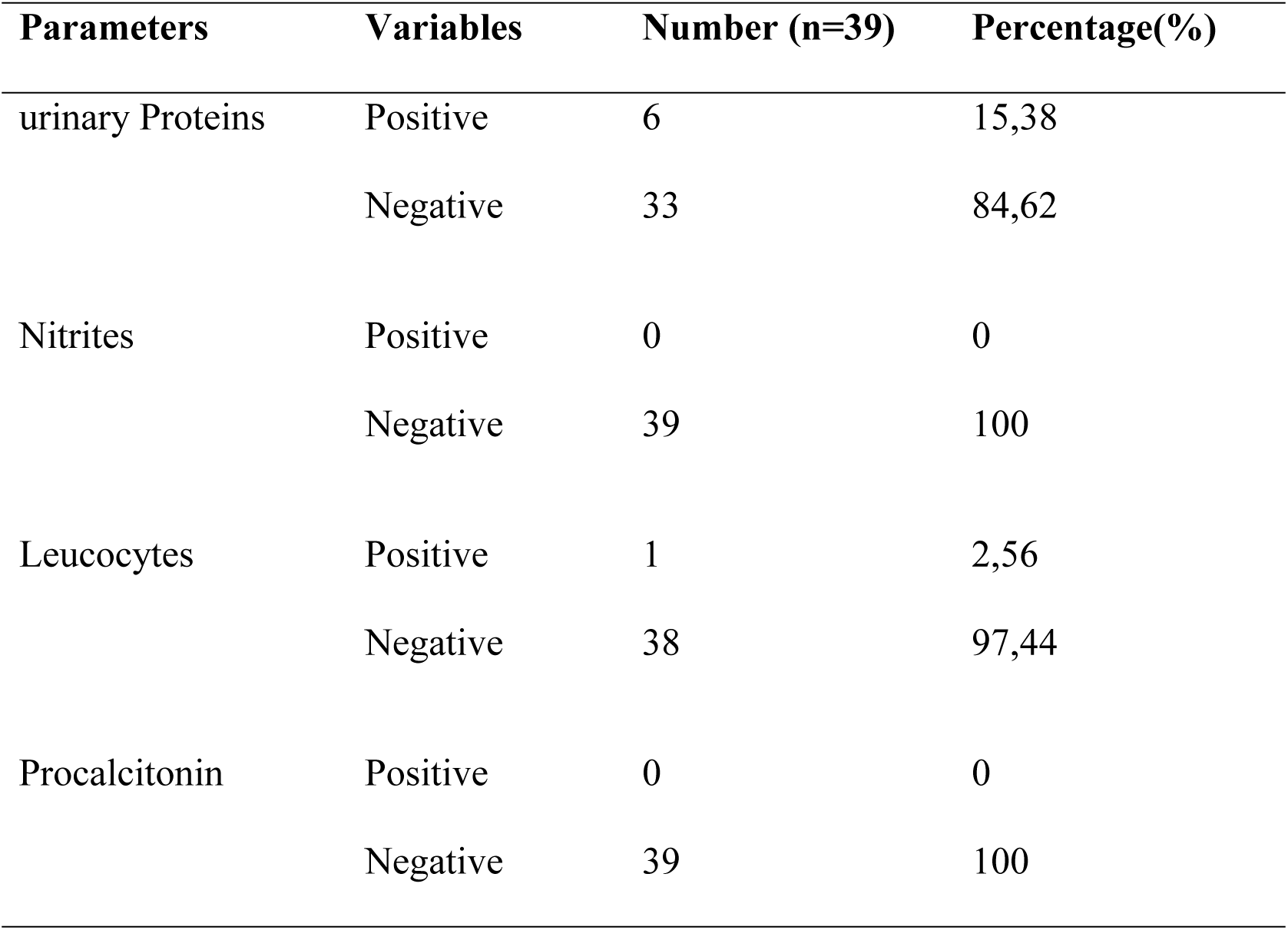
Urine dipstick and procalcitonin results.

| Parameters | Variables | Number (n=39) | Percentage(%) |
| --- | --- | --- | --- |
| urinary Proteins | Positive | 6 | 15,38 |
|  | Negative | 33 | 84,62 |
| Nitrites | Positive | 0 | 0 |
|  | Negative | 39 | 100 |
| Leucocytes | Positive | 1 | 2,56 |
|  | Negative | 38 | 97,44 |
| Procalcitonin | Positive | 0 | 0 |
|  | Negative | 39 | 100 |

### Total serum protein

65% of asymptomatic patients had normal serum proteins and 35% abnormal serum proteins, with a minimum of 48.05 g/L and a maximum of 91.70 g/L. In asymptomatic infected patients, we obtained 80% elevated protidemia compared. 8.33% of symptomatic patients had normal protidemia compared to 91.67% with elevated protidemia. They had a minimum of 73.90 g/L, a maximum of 98.20 g/L and a mean of 88.7408 ± 2.04 g/L; the standard deviation was 7.06 g/L and the median was 86.80 g/L. The characteristics of these different groups are summarised in the figure below.

### Results of serum protein electrophoresis

All the albumin fractions showed a relative decrease in all our patients, with a minimum of 44.90% (for asymptomatic infected) and a maximum of 54.60% (for asymptomatic uninfected). All the symptomatic and infected patients had abnormal α-1 globulin fractions. 50% had a decrease in the α-2 globulin fraction. β-1and β-2 globulin fractions were normal for all the patients. All gamma globulin fractions were higher in all patients, with a mean of 26.25% ± 2.49% and extremes of 21.7 and 37%.

## Discussion

This study aimed to determine the biochemical markers associated with filariasis in the hospital at the Moungo Division.

The study population was highly dominated by females. The sex ratio (F/M) was 2.66. In most articles in the literature, the population is dominated by males [23–25].

35% of the population have microfilaria and 65% were not infected. These data are higher than that of Wandji *et al.,* [26] on top of these changes, these authors did not find *Wuchereria bancrofti*, in their studies sites in Cameroon. Moreover, these results are higher than that of Nana-Djeunga *et al.*, [27] who drew the map *Wuchereria bancrofti* throughout Cameroon and found that the prevalence of detection of microfilaria was between 0 and 1% in the same study area. Out of Cameroon, these data are lower compared to that of Lamontellerie *et al.*, [28] that found in Burkina Faso, 85.4% of the same filariasis. More specifically, 18,18% of our study population was infected but still asymptomatic. This goes in the same line with the report from Nana-Djeunga *et al.*, [27]. These authors found that even though the filarial load was low in many regions of Cameroon, the global circulating filarial antigen was at around 20% of his study population. 21.81% were symptomatic and had microfilaria in their blood. From this positive population, Onchocerca (18.2%), *Mansonella perstans* (54,54%), *Loa loa* (22.72%) and *Wuchereria bancrofti* (4.54%) were detected. This does not correspond to the report from Wandji *et al*., [26] who found that *Loa loa* was the most present in localities of the East Region of Cameroon followed by *Mansonella perstans*. In Burkina Faso, Lamontellerie *et al.*, [28] found onchocerca (32,4%), *Wuchereria bancrofti* (13,2%) and *Mansonella perstans* (33.8%). Moreover, Kuété *et al.*, [37] found that *O. volvulus* (26,38%), *Loa loa* (5,53%), and *Mansonella perstans* (4,7%) were found in Ntui in Cameroon. These differences observed in different regions and periods in Cameroon might be due to the massive drug administration done by the Cameroon state against lymphatic filariasis.

Taking into consideration the time after the massive drug administration campaign, 33.33% were asymptomatic and the same size were symptomatic two months after the campaign. 15.38% were still symptomatic and infected one year after the campaign. After two years after the campaign, 40% of the patients were symptomatic and infected. These findings are different from those of Camara *et al.,* [29] at Sikasso region, Mali. These authors found that 9 months after the massive drug administration campaign, 2.33% of the population was still symptomatic and infected. This difference might be associated with the re-introduction of the disease from the neighbouring area where the disease is present [27] and mainly by the crisis in North West and South West regions of Cameroon that has led to a massive displacement of the population.

No microfilaria was detected in the urine of the patients. This is in accordance with the literature[30,31]. These research groups found out that the search for microfilaria in urine is important for diagnosis but the detection remains difficult and is mostly associated with another clinical feature. In addition, the results obtained from the urinary stick do not correlate significantly with the presence of microfilaria in the blood. Therefore, the presence of leukocytes in urine was probably associated with urinary infection by pathogens other than lymphatic filaria.

Procalcitonin is a valuable tool to assess the inflammatory response of the organism due to the infection by microorganisms. It is more specific with bacterial infections and slightly interesting for the diagnosis of parasitic diseases[32–37]. The level of procalcitonin does not depict the presence of any infection and concerning the abundant literature on that particular test[32–37], the results confirm that our participants do not suffer from bacteraemia or fungemia.

The lymphatic system is involved in several functions, including the regulation of blood volume through the transport and return of plasma proteins that have escaped from the bloodstream into the blood, and this function could be altered by the presence of microfilariae in the lymphatic system [38]. Therefore, monitoring proteinemia and proteinuria can be of help in the early diagnosis of filariasis[39]. In this study, 91.67% of symptomatic infected and non-infected patients have abnormal proteinemia. This falls in the same line with the observation by Nana-Djeunga *et al.*, [27] who found that the biochemical parameter goes down slowly after clearance of the filaria in the body. We also observed that the proteinemia was 80% higher in asymptomatic infected patients in contrast, the asymptomatic non-infected patients had their protidemia 35% higher than the normal range. We did not find in the literature authors that analysed the protidemia of the uninfected population from that particular rural area.

On the other hand, the albumin fraction was globally low in all study populations, it was lower in infected asymptomatic and symptomatic patients. Regarding the other protein fraction, we found that gamma globulin level was high in all infected patients. This result goes in the same line with that of Moreau *et al.,* in France who found that *Wuchereria Brancrofti*, *Loa loa* and *Mansonella Perstans* infection are correlated with an elevation of immunoglobulin G [40].

## Conclusion

This study highlights the presence of *Onchocerca volvulus*, *Loa loa*, *Wuchereria brancrofti* and *Mansonella perstans* in the Moungo division of the Littoral region. It also highlights that the presence of microfilaria is poorly correlated to proteinemia and correlated to the albumin and gamma globulin levels of blood.

## Data Availability

the data are available at the FMSP-Udo

## Acknowledgements

The authors are grateful to all participants for their collaboration The authors are grateful to all the heads of health care centres in the Moungo division for their support during data collection. Also, the authors would like to thank the Director of the gyneco- obstetric Hospital of Douala for the support during data analysis.

## References

1. Association Française des Enseignants de Parasitologie et de Mycologie (ANOFEL). Filarioses humaines. Univ Médicale Virtuelle Francoph. 2014;1–22.

2. Organisation mondiale de la Santé (OMS). Manuel à l’intention des programmes nationaux : Programme mondial pour l’élimination de la la filariose lymphatique. 107p. mise à jour 2018. 2018;10–22.

3. HAS. Actualisation de la nomenclature des actes de biologie médicale pour le diagnostic et le suivi des filarioses. Haute Autorité de santé. 2018;1–85.

4. Moyou R.S., Ouambe M.A., Fon E. BJ. Enquete Sur La Filariose Lymphatique Dans Sept Villages Du District. Med Trop. 2003;63:583–6.

5. Laboratoire du Centre Pasteur. filariose lymphatique au Cameroun : état des connaissances. Bull liais doc OCEAC1999. 1987;32(4):(15 mars 2008).

6. Valérie D. Onchocercose _ définition, causes et prévention. passeport santé/maladies.

7. Adrian H., S. P. Élimination De L’Onchocercose Et De La Filariose Lymphatique. Numéro. 2015;12:14–6.

8. Tongue R. Une nouvelle stratégie pour éliminer les filaires au Cameroun - Médiaterre.

9. Nizeyimana JB, Mabée JM, Zongo JM. Étude Des Conditions De Depistage Clinique Et Les Effets Psycho Socioeconomique Des Personnes Porteuses De L’Elephantiasis Des Jambes. Cas De L’Arrondissement De Guidiguis Cas De La Region De L’Extreme-Nord Cameroun De 2017-2020. Vol. 26, RUFSO Journal of Social Sciences and Engineering. 2021.

10. Kamgno J, Boussinesq M. Hyperendémicité de la loase dans la plaine Tikar, région de savane arbustive du Cameroun. Bull la Soc Pathol Exot. 2001;94(4):342–6.

11. Collomb H, Miletto G. Filariose lymphatique. Vol. 6, La Revue du praticien. 1956. p. 839–50.

12. Pierre A. BAG. Filarioses lymphatiques: actualités 2021. :1–8.

13. Laurent Y., Christian L. JMH. Trente ans de lutte contre l’onchocercose en Afrique de l’Ouest. Traitements larvicides et protection de l’environnement. Trente ans de lutte contre l’onchocercose en Afrique de l’Ouest. Traitements larvicides et protection de l’environnement. 2003.

14. La France au Cameroun. Comprendre la filariose à Loa Loa au Congo Brazzaville et au Cameroun : Lancement des préparatifs du projet europeen Morlo. Ambassade de France à Yaoundé.

15. OMS. RAPPORT TECHNIQUE ANNUEL DES ACTIVITES DE LUTTE CONTRE LES MALADIES TROPICALES NEGLIGEES 2017 (Additif : PLAN D ACTION ANNUEL 2018). 2017;1–48.

16. Kamgno J., Tsague D., Gardon J. BM. Données complémentaires sur l’endémie à Loa loa dans la Province du Centre du Cameroun- fdi_010028375- Horizon. 2001 p. Bulletin de Liaison et de Documentation de l’OCEAC.

17. CDC. CDC - Onchocerciasis - Resources for Health Professionals. CDC - Onchocerciasis - Resources for Health Professionals. 2021. p. Online.

18. Kamgno J, Nana-Djeunga HC, Pion SD, Chesnais CB, Klion AD, Mackenzie CD, et al. Operationalization of the test and not treat strategy to accelerate the elimination of onchocerciasis and lymphatic filariasis in Central Africa. Int Health. 2018;10(March):i49–53.

19. Mfoumou B. Le Cameroun adopte la stratégie _Test and Treat_ pour combattre l’onchocercose. Cameroon Radio Television.

20. Lepori A sophie. L ’ onchocercose : données actuelles et nouvel horizon thérapeutique : le rôle de la doxycycline dans le traitement de l ’ onchocercose To cite this version : HAL Id : hal-01732568 soutenance et mis à disposition de l ’ ensemble de la Contact : ddoc-these. 2018;107.

21. Djako A. Epidémiologie spatiale des maladies tropicales négligées: cas de Maroua - extrème nord, Cameroun. :30.

22. BIOFORMA. Cahier de formation Biologie médicale N° 23 - Parasites sanguins. 2001. 357 p.

23. Et S, Recherche DELA, Sciences UDES, Techniques DES, Des ET, Bamako TDE. These de medecine. 2020;

24. Amaechi EC, Ohaeri CC, Ukpai OM, Nwachukwu PC, Ukoha U. Asian Pacific Journal of Tropical Disease. Asian Pacific J Trop Dis [Internet]. 2016;6(9):709–13. Available from: 10.1016/S2222-1808(16)61114-3

25. Soc TR, Med T. The impact of Loa loa microfilaraemia on research subject retention during a whole sporozoite malaria vaccine trial in Equatorial Guinea. 2022;(April):745–9.

26. Wanji S, Esum ME, Njouendou AJ, Mbeng A, Ndongmo PWC, Abong RA, et al. Mapping of lymphatic filariasis in loiasis areas : A new strategy shows no evidence for Wuchereria bancrofti endemicity in Cameroon. PLoS Negl Trop Dis. 2019;13(3):1–15.

27. Nana-djeunga HC, Tchatchueng-mbougua JB, Bopda J. Mapping of Bancroftian Filariasis in Cameroon : Prospects for Elimination. PLoS Negl Trop Dis. 2015;9(9):1– 19.

28. Lamontellerie M. Résultats d’enquêtes sur les filarioses dans l’Ouest de la Haute-Volta (Cercle de Banfora). Ann Parasitol Hum Comparée. 1972;47(6):783–838.

29. Camara MM. Dédicaces. 2012;1–46.

30. Mathison BA, Couturier MR, Pritt BS. Diagnostic identification and differentiation of microfilariae. J Clin Microbiol. 2019;57(10):1–13.

31. Sarangi J, Arava S, Kumar H. Microfilaria in urine cytology: Report of three cases with review of literature. Diagn Cytopathol. 2020;48(7):675–8.

32. Hausfater P. Procalcitonine et infection. Ann Fr Med d’Urgence. 2011;1(3):206–12.

33. Samsudin I, Vasikaran SD. Clinical utility and measurement of procalcitonin. Clin Biochem Rev. 2017;38(2):59–68.

34. de Carvalho FT, Rabello Filho R, Bulgarelli L, Serpa Neto A, Deliberato RO. Procalcitonin as a Diagnostic, Therapeutic, and Prognostic Tool: a Critical Review. Curr Treat Options Infect Dis. 2019;11(1):1–11.

35. Lee H. Procalcitonin as a biomarker of infectious diseases. Korean J Intern Med. 2013;28(3):285–91.

36. Wright CA, Path FRC, Burg M Van Der, Ph D, Geiger D, Sc M. Diagnosing Mycobacterial Lymphadenitis in Children Using Fine Needle Aspiration Biopsy : Cytomorphology, ZN Staining and Autofluorescence — Making More of Less. Diagn Cytopathol. 2008;36(4):245–51.

37. Azzini AM, Dorizzi RM, Sette P, Vecchi M, Coledan I, Righi E, et al. A 2020 review on the role of procalcitonin in different clinical settings: an update conducted with the tools of the Evidence Based Laboratory Medicine. Ann Transl Med. 2020;8(9):610– 610.

38. Constantine M, Fili V, Biologiques S. Cours L3 IMMUNOLOGIE (Pr TEBIBEL . S) Matière / Hématologie cellulaire et hématopoïèse Chapitre 1 Le milieu intérieur. :1–11.

39. Durand G. Jean-Louis Beaudeux, Biochimie médicale: Marqueurs actuels et perspectives, Chapitre 7: Les marqueurs biochimiques de l’inflamation, P 99-112; 2e édition, Médecine Sciences Publications, Lavoisier.

40. Moreau J, Cuzon G, Pichon G, Outin-Fabre D, J. Lagraulet. Les protbines sbriques du filarien lymphatique a wuchereria bancrofti var. Pacifica. Btude blectrophori∼tique et dosage immunochimique des immunoglobulines a, m, g et e. Bull LA SOCIlTE Pathol Exot. 1972;456–63.

